# Population based targeted sequencing of 54 candidate genes identifies *PALB2* as a susceptibility gene for high grade serous ovarian cancer

**DOI:** 10.1101/19011924

**Authors:** Honglin Song, Ed Dicks, Jonathan P. Tyrer, Maria Intermaggio, Georgia Chenevix-Trench, David D Bowtell, Nadia Traficante, AOCS Group, James D. Brenton, Teodora Goranova, Karen Hosking, Anna Piskorz, Elke Van Oudenhove, Jennifer Anne Doherty, Holly R. Harris, Mary Anne Rossing, Matthias Dürst, Thilo Dörk, Natalia V. Bogdanova, Francesmary Modugno, Kirsten B. Moysich, Kunle Odunsi, Roberta B. Ness, Beth Y. Karlan, Jenny Lester, Allan Jensen, Susanne K. Kjaer, Estrid Høgdall, Ian Campbell, Conxi Lazaro, Miquel Angel Pujana, Julie M. Cunningham, Robert A. Vierkant, Stacey J. Winham, Michelle A.T. Hildebrandt, Chad Huff, Donghui Li, Xifeng Wu, Yao Yu, Jennifer B. Permuth, Douglas A. Levine, Joellen M. Schildkraut, Marjorie J. Riggan, Andrew Berchuck, Penelope M. Webb, OPAL Study Group, Cezary Cybulski, Jacek Gronwald, Anna Jakubowska, Jan Lubiński, Jennifer Alsop, Patricia A. Harrington, Isaac Chan, Usha Menon, Celeste L. Pearce, Anna H. Wu, Anna de Fazio, Catherine J. Kennedy, Ellen L. Goode, Susan J. Ramus, Simon A. Gayther, Paul D.P. Pharoah

## Abstract

**Purpose:** The known EOC susceptibility genes account for less than 50% of the heritable risk of ovarian cancer suggesting other susceptibility genes exist. The aim of this study was to evaluate the contribution to ovarian cancer susceptibility of rare deleterious germline variants in a set of candidate genes.

**Methods:** We sequenced the coding region of 54 candidate genes in 6385 invasive EOC cases and 6115 controls of broad European ancestry. Genes with an increased frequency of putative deleterious variants in cases verses controls were further examined in an independent set of 14,146 EOC cases and 28,661 controls from the ovarian cancer association consortium and the UK Biobank. For each gene, we estimated the EOC risks and evaluated associations between germline variant status and clinical characteristics.

**Results:** The odds ratios (OR) associated for high-grade serous ovarian cancer were 3.01 for *PALB2* (95% CI 1.59 – 5.68; P = 0.00068), 1.99 for *POLK* (95% CI 1.15 – 3.43; P = 0.014), and 4.07 for *SLX4* (95% CI 1.34-12.4; P = 0.013). Deleterious mutations in *FBXO10* were associated with a reduced risk of disease (OR 0.27, 95% CI 0.07 −1.00, P=0.049). However, based on the Bayes false discovery probability, only the association for *PALB2* in high-grade serous ovarian cancer is likely to represent a true positive.

**Conclusions:** We have found strong evidence that carriers of *PALB2* deleterious mutations are at increased risk of high-grade serous ovarian cancer. Whether the magnitude of risk is sufficiently high to warrant the inclusion of *PALB2* in cancer gene panels for ovarian cancer risk testing is unclear; much larger sample sizes will be needed to provide sufficiently precise estimates for clinical counselling.

## INTRODUCTION

Rare, predicted deleterious variants in multiple genes have been shown to be associated with a moderate to high risk of epithelial ovarian cancer (EOC). These include the DNA double stand break repair genes *BRCA1*^[1]^, *BRCA*^*[2]*^, *BRIP1*^*[3]*^, *RAD51C, and RAD51*^*[4]*^, and the mismatch repair genes *MSH2* and *MSH6*^[5 6]^. *ANKRD11, FANCM* and *POLE* have recently been reported as possible susceptibility genes^[7 8]^. Multiple common variants conferring weaker risk effects have also been identified^[9-16]^, some of which modify EOC risk in carriers of more highly penetrant gene mutations[17 18].

EOC is heterogeneous with five main histotypes: High-grade serous (HGSOC), low-grade serous (LGSC), endometrioid, clear cell and mucinous ovarian cancer. These have different clinical characteristics and outcomes and are characterized by different germline and somatic genetic changes that result in the perturbation of different molecular pathways. For example, germline mutations in DNA double break repair genes predispose to HGSOC while germline mutations in mismatch repair genes increase risk of the endometrioid and clear cell histotypes[6].

The known susceptibility alleles account for less than 50% of the excess familial risk of ovarian cancer, suggesting that other susceptibility genes and alleles exist [14]. The unexplained genetic component of risk is likely to be made up of a combination of common genetic variants conferring weak effects and uncommon alleles conferring weak to moderate relative risks (less than 10-fold).

The aim of this study was to identify additional ovarian cancer susceptibility genes using case-control sequencing of candidate genes identified through various approaches including their known function in pathways that are associated with ovarian cancer development, and from whole exome sequencing studies of ovarian cancer cases that have identified putative deleterious mutations in genes not previously evaluated for EOC risk.

## MATERIAL AND METHODS

### Selection of candidate genes

#### Genes based on known biological function

As several EOC susceptibility genes are involved in DNA double-strand break repair and Fanconi anemia (FA) [8], we selected genes involved in these pathways. FA is a rare genetic disease characterized by chromosomal instability, hypersensitivity to DNA crosslinking agents, defective DNA repair, severe bone marrow failure, cancer susceptibility and many congenital defects. To date, more than 20 FA genes have been identified. We selected eight FA genes not previously studied in ovarian cancer: *FANCA, FANCB, FANCC, FANCD2, FANCE, FANCG, FANCI, FANCP (SLX4)*. We also included *FANCN (PALB2)*, which has been studied previously in ovarian cancer [3 19-21] but its association with EOC risk is equivocal. Eight candidate genes involved in other aspects of DNA repair were also included: *ALKBH3, CHEK2, GTF2H4, POLE, POLK, RDM1, REV3L*, and *XRCC1*.

#### Genes from whole exome sequencing studies (WES)

Twelve genes *(BUB1B, C5orf28, C6, DNAJB4, EXO1, LIG4, MKNK2, MMRN1, PARP1, RAD52, SMC1A* and *SNRNP200)* were selected from WES analysis of EOC cases where putative deleterious (truncating) mutations were identified at a greater frequency in cases compared to publicly available WES data from controls reported by the NHLBI GO Exome Sequencing Project (ESP) and The Exome Aggregation Consortium (ExAC) databases (http://exac.broadinstitute.org). Germline WES data for EOC cases were available for 412 HGSOC cases from the Cancer Genome Atlas (TCGA) ovarian cancer study; 513 ovarian cancer cases from an Australian case series; 97 familial non-*BRCA1/BRCA2* ovarian cancer cases from Gilda Radner Familial Ovarian Cancer Registry (GRR); and 54 ovarian cancer cases from the UK Familial Ovarian Cancer Registry (UKR).

Four genes from these WES studies (*GANC, KNTC1, PSG6 and UPK2*) were selected because more than one family member diagnosed with ovarian cancer from 10 familial cases carried the same truncating mutation in one of these genes.

Finally, twenty-one genes were selected from analyses of several other unpublished EOC WES studies (personal communications) where the frequency of truncating mutations was greater in cases compared to controls. These genes were *ANAPC2, CNKSR1, DUOX1, FBXO10, NAT10, OSGIN1, PAK4, PHF20L1, PIK3C2G, PTGER3, PTX3, RAD54B, RECQL, RIPK3, RNASEL, SMG5, SPHK1, SULT1C2, UHRF2, WNT5A and ZFHX3*.

### Study subjects

We used case-control data from targeted sequencing, exome and array-based genotyping.

#### Targeted sequencing

We included 5,914 EOC cases and 5,479 controls of European ancestries from nineteen studies - thirteen case-control studies, one familial ovarian cancer study from Poland, two clinical trials and three case-only studies **(Supplementary Table 1)**[13]. HGSOC cases were preferentially plated out for sequencing where possible.

#### Exome sequencing

We extracted data on the 54 candidate genes from 829 case and 913 controls from two ovarian cancer case-control studies (MDA[22-24]and NCO[13]) for which whole exome sequence data were available **(Supplementary Table 1)**.

#### Variants from genotyping array data

For genes that reached nominal significance in the combined analysis of the targeted sequencing and exome sequencing data, we extracted genotypes of any deleterious variants included on the OncoArray and UK Biobank Axiom Array. These two arrays were used to genotype up to 18,936 controls and 13,288 cases from the Ovarian Cancer Association Consortium (OCAC)[14], 9,725 controls and 858 cases from UK Biobank GWAS (https://www.ukbiobank.ac.uk/) respectively. Samples overlapped with the sequencing studies were excluded from the analysis.

All studies had ethics committee approval, and all participants provided informed consent.

### Sequencing methods

Target sequence enrichment followed by sequencing was performed on the coding sequence and splice-sites of ALKBH3, ANAPC2, BUB1B, C5ORF28, C6, CHEK2, CNKSR1, DNAJB4, DUOX1, EXO1, FANCA, FANCB, FANCC, FANCD2, FANCE, FANCG, FANCI, FBXO10, GANC, GTF2H4, KNTC1, LIG4, MKNK2, MMRN1, NAT10, OSGIN1, PAK4, PALB2, PARP1, PHF20L1, PIK3C2G, POLE, POLK, PSG6, PTGER3, PTX3, RAD52, RAD54B, RDM1, RECQL, REV3L, RIPK3, RNASEL, SLX4, SMC1A, SMG5, SNRNP200, SPHK1, SULT1C2, UHRF2, UPK2, WNT5A, XRCC1 and ZFHX3, using 48.48 Fluidigm access arrays as previously described[6]. A total of 1,663 amplicons were designed to cover the 159kb target region. Libraries were sequenced using 150 bp paired-end sequencing on the Illumina HiSeq4000 or HiSeq2500.

Sequencing reads were de-multiplexed then aligned against the human genome reference sequence (hg19) using the Burrows-Wheeler Aligner (BWA) [25]. The Genome Analysis Toolkit (GATK) [26] was used for base quality-score recalibration, local indel realignment and variant calling. Finally, ANNOVAR [27] was used for variant annotation. Variants were called if (1) genotype information was available from a chip genotype for that sample, or (2) the variants were presented in more than one amplicon, or (3) read depth ≥15 and alternate allele frequency ≥40 percent, or (4) read depth ≥100 and alternate allele frequency ≥ 25 percent. These thresholds were defined using the results from sequencing of positive controls with known variants and genotype information from chip array genotyping of overlapping samples.

We excluded 356 cases and 269 controls because <80 percent of the target sequence bases had a read depth of at least 15. The average percentage coverage of the genes at 15X read depth ranged from 64% - 99% **(Supplementary Table 2)**. The mean sequencing depth for these genes ranged from 130 (IQR 104 – 152) to 432 (IQR 364 – 492). Concordance for 111 duplicate pairs was 98% (7384 concordant variants out of total 7572 variants called).

For the exome sequencing, sonication fragmentation was used to fragment DNA samples. Fragments with an average size of 200bp were selected to generate libraries for sequencing. Agilent SureSelect Clinical Research Exome (CRE) v1 was used for exome enrichment and sequencing was performed on an Illumina HiSeq 4000 using 2×150bp paired-end reads. Cutadapt (https://doi.org/10.14806/ej.17.1.200) was used to locate and remove residual adapters in reads. FLASH (Fast Length Adjustment of SHort reads)[28] was used to merge the overlapped paired-end reads into one read, using default parameters. Reference genome alignment and joint genotype calling according to a pipeline described in Yu et al[29]. The coding sequences and splice sites of all 54 genes were extracted. Fifty-three genes with 100% average coverage at 10X were included in the analysis. *GTF2H4* was excluded from the analysis, as the average coverage was only 43%.

Deleterious variants were defined as those predicted to result in protein truncation (frameshift indel, splice site, nonsense mutations and start loss) or predicted to be deleterious and/or likely deleterious by Clinvar[30]. We used the software MaxEntScan to identify splice site variants most likely to affect gene splicing [31] – those with a MaxEntScan score that decreased by more than 40% compared to the reference sequence and having a reference sequence score ≥3. Sequencing alignments were confirmed by visual inspection using the Integrative Genomic Viewer (IGV) [32].

### Statistical methods

#### Risk Estimation and genotype-phenotype analyses

We used a simple burden test for association between deleterious variants and ovarian cancer risk on a gene-by-gene basis. The burden test was based on unconditional logistic regression adjusted for country (Australia, Denmark, German, Poland, the United Kingdom and the USA) and sequencing method (targeted sequencing or exome sequencing). Odds ratios and associated 95 percent confidence intervals (95% CI) were calculated.

#### Missense variant analyses

We also identified multiple rare (minor allele frequency < 1%) missense variants that have an unknown functional effect on the protein. We used the rare admixture likelihood (RAML) burden test[33] to test these variants for association. We classified variants by whether or not they are predicted to have a damaging effect on protein function by 2 out of 3 prediction tools - SIFT (score <0.05)[34], polyphen-2[35] (classified as probably damaging or damaging) and Provean[36] (score<=-2.5). Subjects with a missense variant call rate less than 80 percent and variants with a call rate less than 80 percent or with genotype frequencies inconsistent with Hardy-Weinberg equilibrium (P<10^−5^) were excluded.

## RESULTS

### Germline deleterious mutations in ovarian cancer cases and controls

Sequencing results were available for 6,385 EOC cases and 6,115 controls after quality control analysis. The characteristics of these individuals by study are summarized in **Supplementary Table 1**. Most EOC cases were serous histotype (N= 6,304, 98.7 percent), of which 5,951 were the HGSOC histotype (93.2 percent).

We identified 628 unique, putative-deleterious variants **(Supplementary Table 3)** in 909 HGSOC cases (14.2 percent) and 873 controls (14.3 percent). There was a nominally significant higher frequency of mutations in cases compared to controls for *POLK, PALB2* and *SLX4* and a lower frequency of mutations in cases compared to controls for *FBXO10* **(Table 1)**. The associated odds ratios are shown in **Table 1** – for *POLK, PALB2* and *SLX4* the effect size was slightly larger for HGSOC. The frequency of deleterious variants in the other genes was similar in cases compared to controls **(Supplementary Table 4)**. Given the evidence for association of multiple Fanconi anemia genes with EOC risk we also carried out a burden test to compare the frequency of deleterious variants in any of the eight genes which were not significantly associated with ovarian cancer risks individually (*FANCA, FANCB, FANCC, FANCD2, FANCE, FANCG, FANCI* and *FANCL*). A combined analysis will have greater power if multiple genes were associated but the effect sizes too small to detect individually. There was no significant difference in the frequency of deleterious variants in cases (96/6184, 1.6%) and controls (85/6,089, 1.4%) (P = 0.50).

**Table 1.**
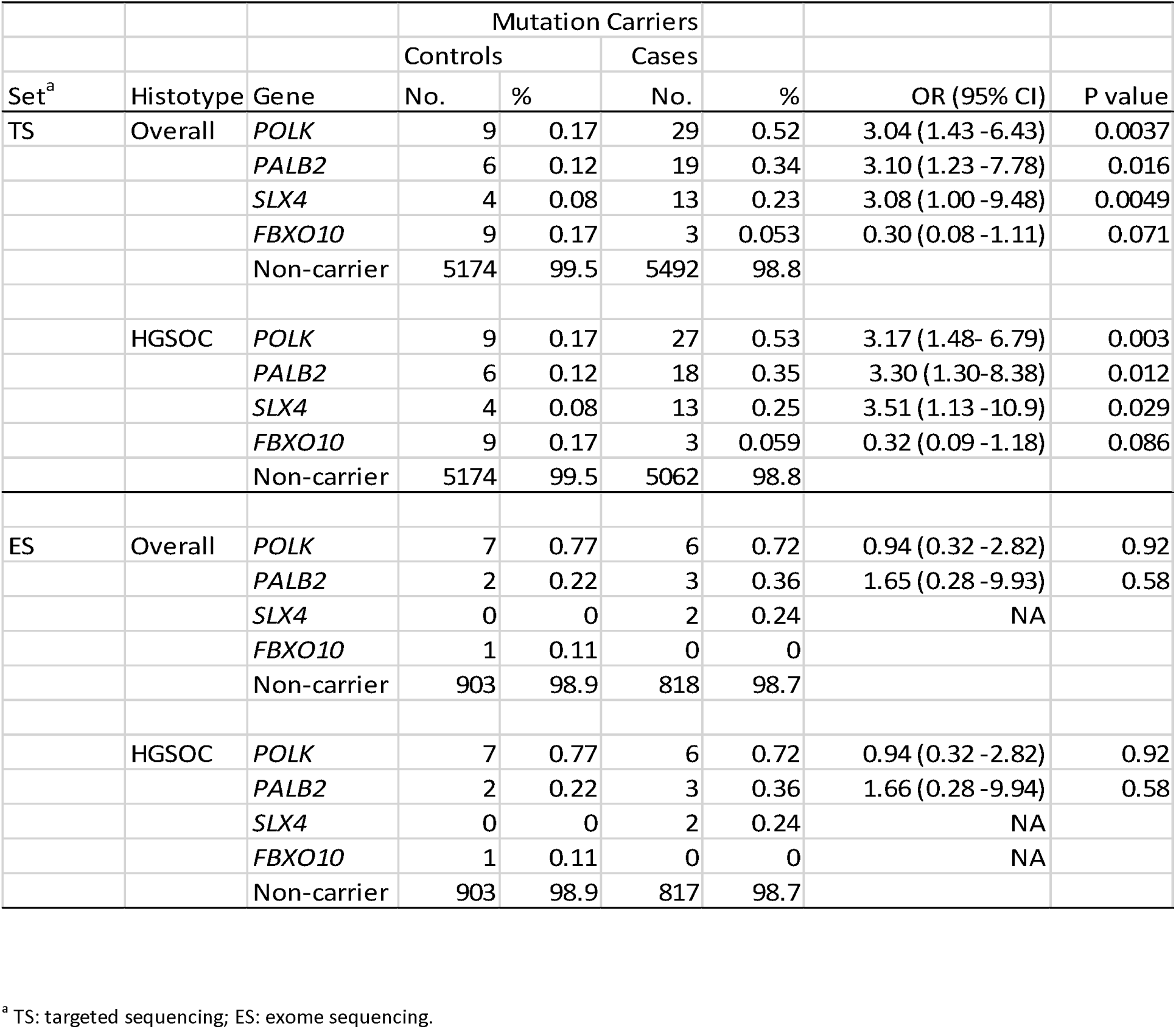
Frequency of mutations and estimated risk of EOC in candidate genes (P<0.05) from targeted sequencing and exome sequencing

### Validation analyses in ovarian cancer case-control studies

We also evaluated risk associations between deleterious variants in *POLK, PALB2*, and *SLX4* with EOC risk based on germline genotyping data for 13,277 EOC cases and 18,930 controls from OCAC and for 858 EOC cases and 9,725 controls and from UK Biobank. For OCAC samples, data were available for six deleterious non-monomorphic variants in PALB2; for UK Biobank samples, data were available for seven *PALB2* and one *POLK* deleterious variants **(Table 2)**.

**Table 2.** Frequency of mutations and estimated risk of EOC in candidate genes for validation chip genotyping data

|  |  |  | Mutation Carriers |  |  |  |  |  |
| --- | --- | --- | --- | --- | --- | --- | --- | --- |
|  |  |  | Controls |  | Cases |  |  |  |
| Set<sup>a</sup> | Histotype | Gene | No. | % | No. | % | OR (95% CI) | P value |
| OCAC | Overall | *PALB2* | 6 | 0.03 | 11 | 0.08 | 2.10 (0.74 -5.94) | 0.16 |
|  |  | Non-carrier | 18930 | 99.97 | 13277 | 99.9 |  |  |
|  | HGSOC | *PALB2* | 6 | 0.03 | 6 | 0.097 | 3.48 (1.10 -11.1) | 0.035 |
|  |  | Non-carrier | 18930 | 99.97 | 6168 | 99.9 |  |  |
| Biobank | Overall | *PALB2* | 11 | 0.11 | 3 | 0.35 | 3.12 (0.87 - 11.2) | 0.081 |
|  |  | *POLK* | 29 | 0.30 | 2 | 0.23 | 0.78 (0.19- 3.29) | 0.74 |
|  |  | *SLX4* | 1 | 0.010 | 0 | 0 | NA |  |
|  |  | Non-carrier | 9684 | 99.6 |  |  |  |  |
|  | HGSOC<sup>b</sup> | *PALB2* | 11 | 0.11 | 1 | 0.28 | 2.49 (0.32 - 19.4) | 0.38 |
|  |  | *POLK* | 29 | 0.30 | 1 | 0.28 | 0.92 (0.12- 6.74) | 0.93 |
|  |  | *SLX4* | 1 | 0.010 | 0 | 0 | NA |  |
|  |  | Non-carrier | 9684 | 99.6 | 361 | 99.4 |  |  |

<sup>a</sup> OCAC: OCAC sample genotype on the OncoArray; Biobank: genotype from UK Biobank Axiom Array.
<sup>b</sup>: Information on tumor grade was not available for UK Biobank cases, all the serous cases in UK Biobank were assumed to be HGSOC.

In OCAC case-control analyses, PALB2 variants showed a non-significant increased risk of EOC (OR 2.10, 95% CI 0.74 −5.94, P=0.16). The strength of this association increased when the analysis was restricted to 6,181 HGSOC cases (OR 3.48, 95% CI 1.10 −11.1, P=0.035). In UK Biobank we observed a weak association for PALB2 mutations with EOC risk (OR 3.12, 95% CI 0.87-11.2, P=0.081). There was no evidence of risk association for mutations in *POLK* and *SLX4* **(Table 2)**.

We then performed a meta-analysis by combining the targeted sequencing, whole exome sequencing and chip genotyping data. Taken together, putative deleterious mutations were associated with increased risk for PALB2 (OR 2.60, 95% CI 1.45 – 4.64; P = 0.0013), *POLK* (OR 1.77, 95% CI 1.07 – 2.93; P = 0.026) and *SLK4* (OR 3.37, 95% CI 1.17-9.70, P=0.024), and decreased risk for *FBXO10* (95%CI 0.07-1.00; P=0.049). After stratifying cases by histological subtype, the estimated risks were higher for HGSOC for *PALB2* (OR 3.01, 95% CI 1.59 – 5.68; P = 0.00068), *POLK* (OR 1.99, 95% CI 1.15 – 3.43; P = 0.014) and *SLK4* (OR 3.92, 95%CI 1.33-11.5; P = 0.013).

We used an approximate Bayes factor to calculate the Bayes false discovery probability (BFDP) described by Wakefield[37] for *PALB2, SLX4, POLK* and *FBXO10* based on several different priors and assuming that the associated risk is unlikely to be greater than an odds ratio of 4 (Table 3). The evidence for association of *PALB2* was strong with a BFDP of less than 15% when the prior on the alternative hypothesis is 0.1. The nominally significant associations for the other three genes are likely to be false positives.

**Table 3.**
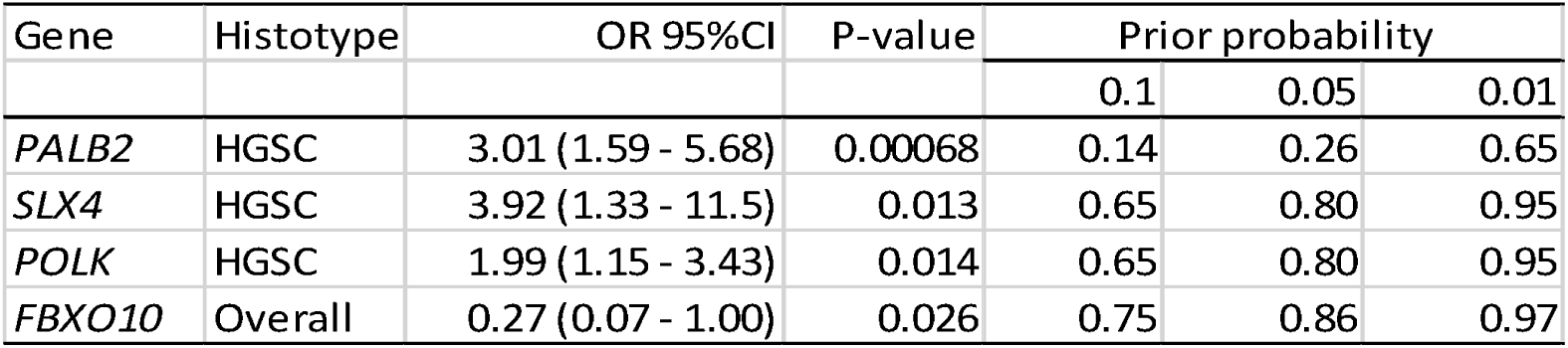
Bayes false discovery probability for the associations reported for *PALB2, SLX4, POLK* and *FBXO10*

| Gene | Histotype | OR 95%CI | P-value | Prior probability |  |  |
| --- | --- | --- | --- | --- | --- | --- |
|  |  |  |  | 0.1 | 0.05 | 0.01 |
| *PALB2* | HGSC | 3.01 (1.59 - 5.68) | 0.00068 | 0.14 | 0.26 | 0.65 |
| *SLX4* | HGSC | 3.92 (1.33 - 11.5) | 0.013 | 0.65 | 0.80 | 0.95 |
| *POLK* | HGSC | 1.99 (1.15 - 3.43) | 0.014 | 0.65 | 0.80 | 0.95 |
| *FBXO10* | Overall | 0.27 (0.07 - 1.00) | 0.026 | 0.75 | 0.86 | 0.97 |

### Predicting the functional impact of missense coding variants

Combining the whole exome and targeted sequencing data, we identified 5,265 unique missense variants with minor allele frequency less than 1% in the 54 genes **(Supplementary Table 6)**. We used the *in silico* software programs SIFT, Polyphen-2 and Provean to evaluate the predicted impact of these variants on protein function for each gene. Of the 5,265 variants, 2,111 were classified as ‘deleterious’ based on at least 2 out of 3 of these classifiers. We found weak evidence for association with increased EOC risk for rare missense variants in *DUOX1* and *PAK4* using burden testing (P= 0.015 and 0.025 respectively); for *DUOX1* the strength of this association improved when the analyses were restricted to the HGSOC histotype (P=0.0061). When we performed the same analyses for 1493 very rare variants (MAF<0.001), we observed significant association for missense variants in *DUOX1* and *FANCE* (P=0.015 and 0.034 respectively).

## DISCUSSION

We have evaluated the association between putative deleterious variants in 54 genes with the risk of HGSOC through a combination of whole exome and targeted sequencing analysis in 5,951 cases and 6,115 controls of broad European ancestries. We found evidence for four genes - *PALB2, POLK, SLX4* and *FBXO10* – associated with HGSOC risk. Association analysis in an additional 13,228 ovarian cancer cases and 28,660 controls genotyped through OCAC and the UK Biobank provided further support for *PALB2* as a *HGSOC* susceptibility gene.

The probability that a genetic association deemed statistically significant is a false positive depends on the prior of the null hypothesis and the power of the study to detect an effect size plausible under the alternative hypothesis. We calculated Wakefield’s BFDP[37] based on several different priors to further evaluate the likelihood that *PALB2, POLK, SLX4* and *FBXO10* are EOC susceptibility genes. If we assume the prior on the alternative to be 1 in 10 or 1 in 20, the BFDPs for the association of deleterious variants in *PALB2* with *HGSOC* are 0.14 and 0.26 respectively. These moderately strong priors are reasonable given the evidence for the association from previously published studies[19]. Two studies have reported nominally significant associations for *PALB2* with odds ratios of OR 4.4 (95% CI 2.1 – 9.1) [19]; and (2.87, 95% CI 1.61 – 4.74)[20]. Kotsopoulos and colleagues reported an increased risk that was not significant (OR, 4.55, 95% CI 0.76 – 27), and, in a subset of the samples included in this study, we also found a non-significant increase in risk (OR 3.2, 95%CI 0.86 – 12)[3].

We lacked the statistical power to identify susceptibility genes conferring relative risks of less than 2 (**Figure 1**). Our use of targeted sequencing and a definition of deleterious variants as those that likely truncate the protein product will have probably underestimated the true prevalence of deleterious variants in these genes. Incomplete coverage of each gene will have missed some small indels and single nucleotide variants. Amplicon based sequencing will also miss large deletions and rearrangements, which are relatively common in some genes [38 39]. Finally, any functional mutations in the non-coding region of these genes will have been missed [40].

**Figure 1:**
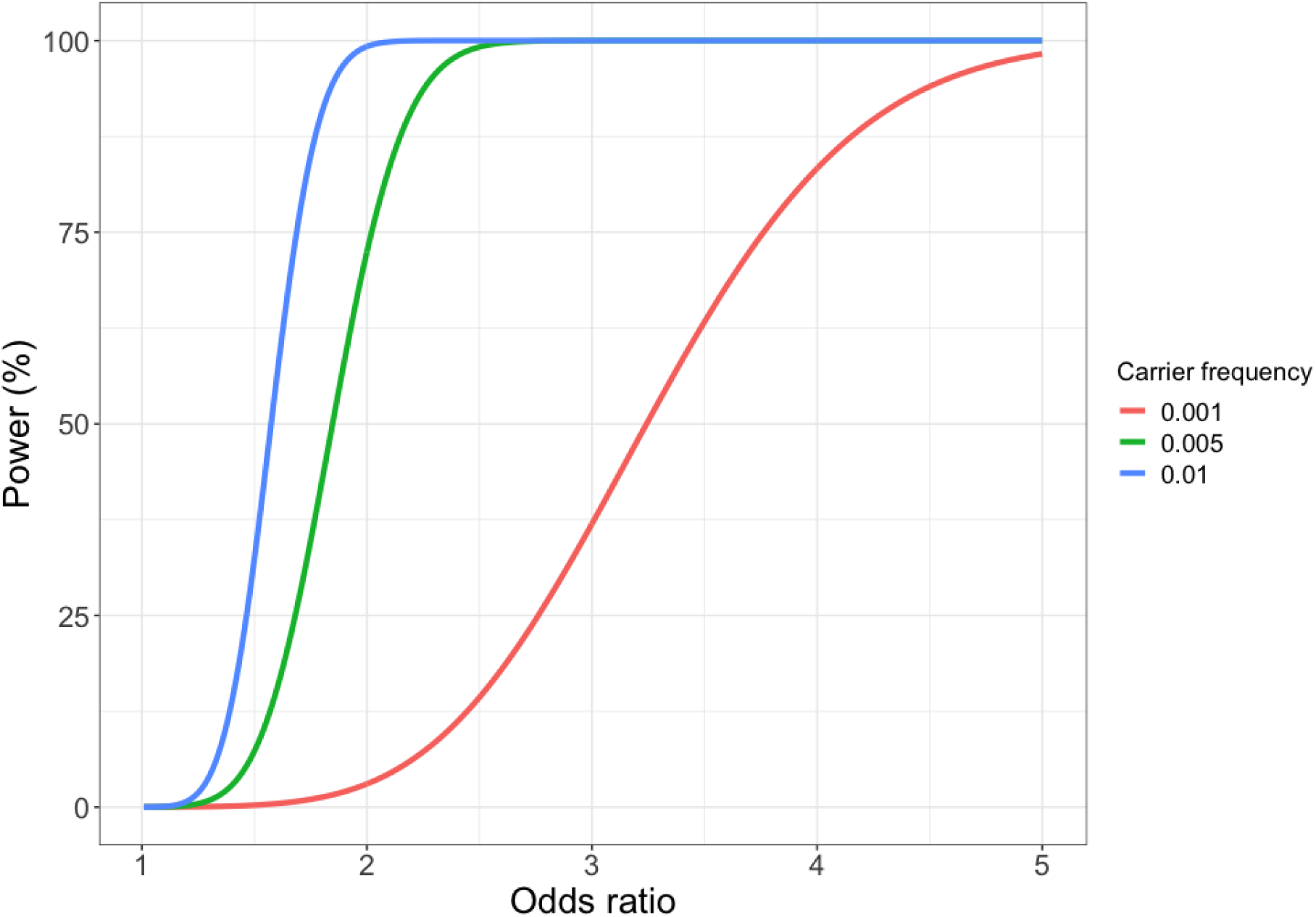
Power to detect association for 5,951 cases and 6,385 controls at a Type I error rate of 0.0001 by deleterious variant carrier frequency and effect size (odds ratio)

Some commercial gene-panel tests for hereditary breast-ovarian cancer already include *PALB2*. However, whether there is clinical utility in testing unaffected women for deleterious mutations in *PALB2* is not clear given the uncertainties in the risk estimates for this gene. There is no consensus over the risk threshold at which preventative surgery should be offered; many cancer genetics clinics in the United Kingdom will refer women if their predicted lifetime risk of epithelial ovarian cancer is greater than 10 percent. Others have suggested that the risk threshold should be lower given the low risk nature of the intervention; prophylactic surgery has been shown to be cost-effective for women at a lifetime risk of 5 percent. Recent updates to the US National Comprehensive Cancer Network (NCCN) Guidelines recommend considering risk reducing salpingo-oophorectomy in carriers of moderate risk genes if the lifetime risk of such mutation carriers exceeds 2.6%. Based on our data and population data for ovarian cancer incidence in England and Wales in 2016, the cumulative risk of ovarian cancer by age 80 for a carrier of a deleterious *PALB2* mutation is 3.2% **(Figure 2)**. Thus, a woman carrying a *PALB2* deleterious mutation would be eligible for prophylactic surgery. However, the confidence intervals for this estimate range from 1.8% to 5.7%. Very large, well-designed case-control studies will be required to provide more precise, unbiased estimates of risk suitable for clinical counselling.

**Figure 2:**
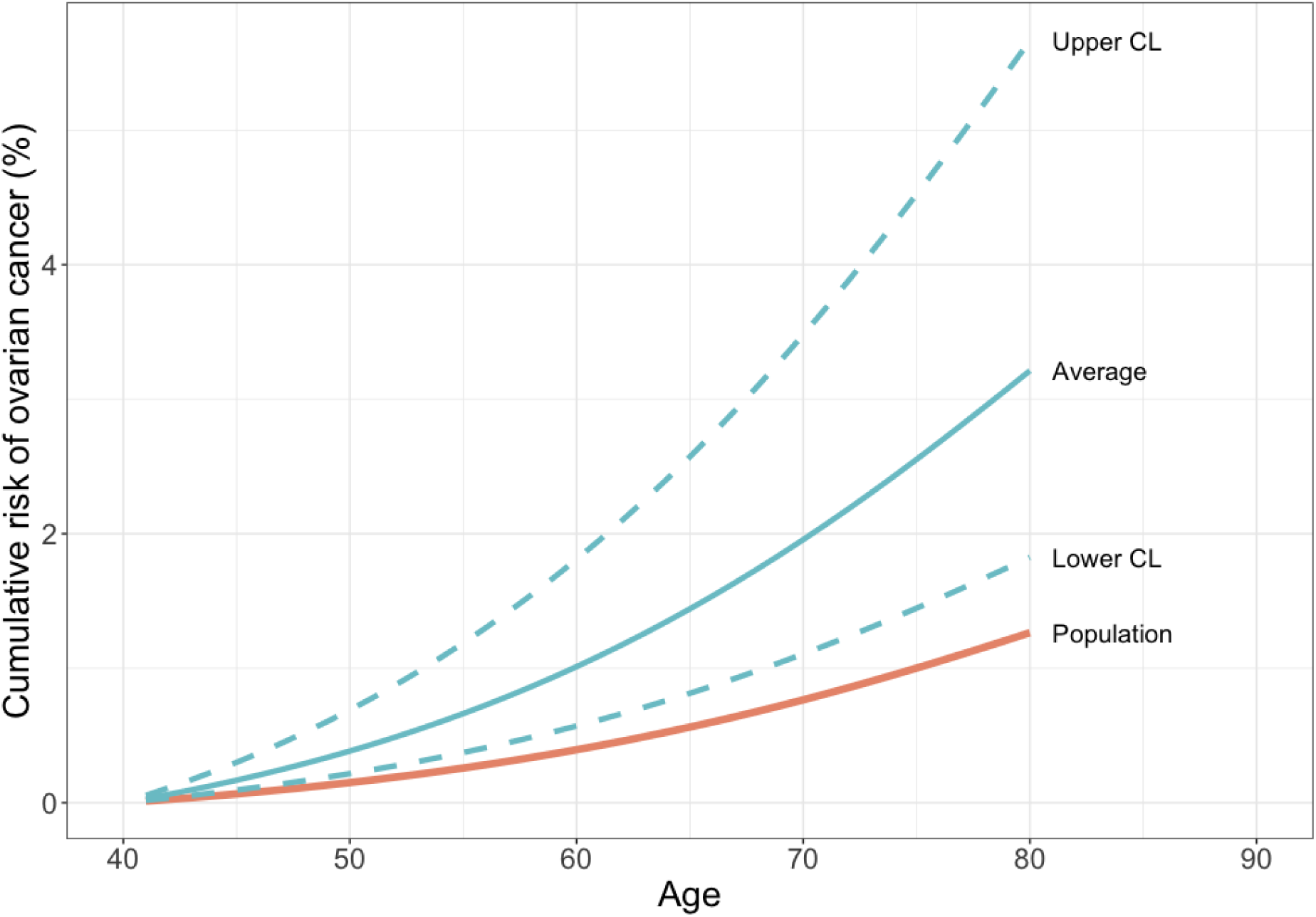
Estimated cumulative risk (%) of ovarian cancer in a *PALB2* deleterious variant carrier compared to population risks for England and Wales, 2016

In summary, we have found relatively strong evidence that deleterious germline mutations in *PALB2* are associated with a moderate increase in the risk of high grade serous ovarian cancer with weak evidence for *POLK, SLX4* and *FBXO10*. Mutations in the other 50 genes we tested do not predispose to HGSOC. This study highlights the importance of large sample sizes needed to obtain risk estimates with the precision necessary for clinical use.

## CONTRIBUTIONS

Honglin Song planned the study, co-ordinated the sequencing, collated the results and drafted the manuscript. Ed Dicks carried out the analysis of the sequencing data. Jonathan P. Tyrer carried out the biostatistical analyses. Georgia Chenevix-Trench, David D Bowtell, Nadia Traficante, James D. Brenton, Teodora Goranova,, Karen Hosking, Anna Piskorz, Elke Van Oudenhove, Jennifer Anne Doherty, Holly R. Harris, Mary Anne Rossing, Matthias Dürst, Thilo Dörk, Natalia V. Bogdanova, Francesmary Modugno, Kirsten B. Moysich, Kunle Odunsi, Roberta B. Ness, Beth Y. Karlan, Jenny Lester, Allan Jensen, Susanne K. Kjaer, Estrid Høgdall, Conxi Lazaro, Miquel Angel Pujana, Julie M. Cunningham, Robert A. Vierkant, Stacey J. Winham, Michelle A.T. Hildebrandt, Chad Huff, Donghui Li, Xifeng Wu, Yao Yu, Jennifer B. Permuth, Douglas A. Levine, Joellen M. Schildkraut, Andrew Berchuck, Penelope M. Webb, Cezary Cybulski, Jacek Gronwald, Anna Jakubowska, Jan Lubiński, Jennifer Alsop, Catherine J. Kennedy, Isaac Chan, Usha Menon, Celeste L. Pearce, Anna H. Wu, Anna deFazio were responsible for data and sample collection for the contributing studies. Marjorie J. Riggan co-ordinated the collection of the OCAC phenotype data. Maria Intermaggio and Patricia A. Harrington carried out the targeted sequencing. Ian Campbell, Ellen L. Goode Susan J. Ramus Simon A. Gayther and Paul D.P. Pharoah planned the study.

## Data Availability

Data are available from authors on request

## FUNDING

American Cancer Society: SIOP-06-258-01-COUN

Cancer Councils of New South Wales, Victoria, Queensland, South Australia and Tasmania and Cancer Foundation of Western Australia: Multi-State Applications 191, 211 and 182

Cancer Institute NSW: 12/RIG/1-17, 15/RIG/1-16

Cancer Research UK: C490/A10119, C490/A10124, C490/A16561, Cambridge Cancer Centre

Kræftens Bekæmpelse: 94 222 52

Medical Research Council: MR_UU_12023

U.S. Department of Health and Human Services, National Institutes of Health, National Cancer Institute: K07CA080668, K22 CA193860, MO1RR000056, P50CA159981, R01CA112523, R01CA122443, P30CA15083, P50CA136393, R01-CA76016, R01CA126841, R01CA178535, R01CA188943, R01CA61107, R01CA87538

R01CA95023

U.S. Department of Health and Human Services, National Institutes of Health, National Center for Advancing Translational Sciences: UL1TR000124

National Health and Medical Research Council of Australia 199600, 310670, 400281, 400413, 628903, APP1025142

National Institutes of Health Research: Cambridge Biomedical Research Centre, University College London Hospitals Biomedical Research Centre

The Eve Appeal: UKOPS Study

U.S. Army Medical Research and Materiel Command: DAMD17-01-1-0729, DAMD17-02-1-0669, DAMD17-02-1-06,

U.S Department of Defense Ovarian Cancer Research Program: W81XWH-07-0449

The University of Cambridge has received salary support in respect of PDPP from the NHS in the East of England through the Clinical Academic Reserve. SAG is a recipient of the Barth Family Chair in Cancer Genetics.

## ACKNOWLEDGMENTS

We thank all the study participants who contributed to this study and all the researchers, clinicians and technical and administrative staff who have made possible this work. In particular, we thank: the clinical and scientific collaborators listed at www.aocstudy.org/ (AOCS) and opalstudy.qimrberghofer.edu.au (OPL).

## CONFLICT OF INTEREST

The authors have no competing interests to declare

## Supplementary Tables legends

Supplementary Table 1. Characteristic of ovarian cancer case-control populations analyzed in this study.

Supplementary Table 2: Target region sequencing coverages and depths for 54 candidate genes.

Supplementary Table 3: Predicted deleterious truncating mutations identified in 54 candidate genes in ovarian cancer case control studies.

Supplementary Table 4: Frequency of mutations and estimated risk of EOC risks in candidate genes (P_overall ≥ 0.05).

Supplementary Table 5: Clinical-pathological characteristics associated with gene carrier status.

Supplementary Table 6: Number of uncommon missense variants (MAF<0.01) detected in cases and controls by gene.

## Notes

### Competing Interest Statement

The authors have declared no competing interest.

### Funding Statement

This research has been supported by NIH/NCI grants R01CA178535 and R01CA188943. The University of Cambridge has received salary support in respect of PDPP from the NHS in the East of England through the Clinical Academic Reserve. SAG is a recipient of the Barth Family Chair in Cancer Genetics. Holly Harris (HRH) is supported by the National Cancer Institute, National Institutes of Health (K22 CA193860). Dr. Beth Karlan is funded by the American Cancer Society Early Detection Professorship (SIOP-06-258-01-COUN) and the National Center for Advancing Translational Sciences (NCATS), Grant UL1TR000124.

